# A Gut-Specific Bispecific Combining MAdCAM-1 Blockade and IL-22 Signaling to Halt T-Cell Inflammation and Promote Mucosal Restoration

**DOI:** 10.64898/2026.08.07.26359969

**Authors:** Juan Diego Sanchez Vasquez, Amanda Sparkes, Navin Asokumar, Jaclyn C. Law, Jean Gariépy

**Affiliations:** Physical Sciences, Sunnybrook Research Institute, Toronto, Canada; Department of Medical Biophysics, University of Toronto, Toronto, Canada; Department of Pharmaceutical Sciences, University of Toronto, Toronto, Canada

## Abstract

Inflammatory bowel disease (IBD) is a heterogeneous chronic disease driven by dysregulated mucosal immunity and impaired epithelial barrier function. Although biologics have improved disease management, they are frequently associated with systemic immunosuppression and adverse effects, highlighting the need for localized therapeutic strategies that both control inflammation and promote tissue repair. Here, we developed a protein bispecific termed 7A2-IgG4-IL22, composed of a human IgG4- Fc domain displaying an antagonistic anti-human MAdCAM-1 single chain (sc)-Fv and a human interleukin (IL-)22.

The anti-MAdCAM-1 scFv retained the functional activity of the parental monoclonal antibody, inhibiting T cell activation, expansion and differentiation from naïve precursors. Blockade of the MAdCAM-1 signaling axis also reduced production of pro-inflammatory cytokines relevant to IBD pathogenesis, including IFN and TNF . On the epithelial side, the IL-22 cargo induces robust signaling in epithelial cells, promoting the expression of IL-22 response genes associated with antimicrobial defense, mucosal homeostasis, as well as *IL-10* and *CXCL1* expression. This effect contributes to immune cell trafficking to the intestinal mucosa. Together, this bispecific provides a localized dual-mechanism strategy for restoring intestinal immune homeostasis.

## Introduction

Inflammatory bowel disease (IBD) refers to a heterogeneous group of lifelong chronic diseases that entail relapsing episodes of intestinal inflammation comprising of two major subtypes: Crohn’s disease (CD) and Ulcerative colitis (UC) based on disease location symptoms and histological characteristics (Monteleone et al., 2025; Saez et al., 2023). CD can affect any part of the gastrointestinal (GI) tract and across all layers of the intestinal wall, especially affecting the small intestine and colon, and is often associated with granuloma formation. In contrast, UC is characterized by inflammation of the mucosal compartment with abnormal activation of the immune system and extends throughout the colon. Predominant symptoms in IBD include diarrhea, abdominal pain and rectal bleeding (Monteleone et al., 2025; Saez et al., 2023; Tavakoli et al., 2021; Yue et al., 2024). IBD exact etiology remains unknown, but multiple lines of evidence suggest that IBD is a multifactorial disease, involving an inappropriate immune response within a genetically susceptible host to environmental factors (Monteleone et al., 2025; Ruan et al., 2025; Saez et al., 2023).

Although the incidence of IBD has stabilized in many western countries, the number of cases remain high, especially in the USA, where the cost of treating IBD patients now exceeds 10 billion dollars per year and is expected to increase (Ruan et al., 2025). Due to the heterogeneity and complexity of the disease there is no definite cure for IBD. Current lines of therapy, including biologics, corticosteroids, aminosalicylates (ASA), and immunosuppressants can induce remission, but many patients remain at high risk of relapse. However, the systemic distribution of such treatments lead to adverse effects such as hepato- and cardiorenal toxicity. In the case of biologics, not all IBD patients are primary responders, and their prolonged use can lead to immunogenicity (Yeshi et al., 2024). Therefore, new therapeutic agents are urgently needed.

The GI tract represents the largest and most complex immune organ as it must maintain balance between responding to pathogens while tolerating luminal contents. However, during inflammation, dynamic events regulating local immune responses can change, leading to the increased migration of lymphocytes into the intestinal mucosa, triggering the aberrant activation of lymphocytes that promote intestinal damage through the excessive release of inflammatory cytokines and chemokines (Yue et al., 2024). Both CD4^+^ and CD8^+^ T cells, despite having distinct profiles, are enriched in lesioned intestinal tissue of IBD patients and have been shown to produce interferon gamma (IFN) and tumor necrosis factor alpha (TNF), cytokines that can lead to mucosal barrier defects and worsening disease prognosis (Funderburg et al., 2013; Imam et al., 2018; Souza et al., 2023).

Mucosal Addressin-Cell Adhesion Molecule 1 (MAdCAM-1) is a cell surface receptor that is mainly expressed by postcapillary venules in the small and large intestinal lamina propria. Importantly, its expression increases within inflamed areas of the gut in IBD patients (Ohtani et al., 2002; Souza et al., 1999; Uchiyama et al., 2023). MAdCAM-1 regulates lymphocyte trafficking into the gut by binding 4 7 integrin on T cells which delivers a co-stimulatory signal that promotes their activation (Briskin et al., 1997; Girard et al., 2024; Grant et al., 2001; Lehnert et al., 1998; Tan et al., 1998; Vimonpatranon et al., 2023). In the inflamed intestine, anti- 4 7 integrin biologics targeting this axis are used in the clinic to reduce lymphocyte trafficking and activation.

For instance, vedolizumab, an approved anti- α 4β7 integrin antibody has been shown to display a higher efficacy profile relative to anti-TNF biologics (infliximab), and IL-12/23 inhibitors (guselkumab and ustekinumab) (Kelly and Long, 2024;Chu et al., 2023). Still, as 4 7 integrin is expressed in circulating lymphocytes, off-target effects remain as integrin 4 7 also interacts with VCAM-1 and increases the risk of progressive multifocal leukoencephalopathy (PML) (D’Haens et al., 2018; Wang et al., 2018; Yeshi et al., 2024). An alternate approach to target this axis would be to block MAdCAM-1 from binding 4 7 integrin. Specifically, the commercially developed anti-MAdCAM-1 antibody ontamalimab, binds and targets human MAdCAM-1. However, despite promising clinical results, its development was suspended (“Update of the Financial Impact of the European Commission’s Decision to Release Takeda from Commitment to Divest Shire’s Pipeline Compound SHP647,” 2020). We have recently developed several monoclonal antibodies that binds and block human MAdCAM-1, preventing T cell homing and activation with its expression being localized to the human intestinal tract (Sparkes et al., 2025). Using one such antagonistic MAdCAM-1 antibody (7A2), we have developed a gut-targeted bispecific where a therapeutic 7A2 single chain (sc)Fv was assembled on a human IgG4 scaffold with another therapeutic cargo, namely human interleukin (IL)-22, to provide dual immunomodulatory functions. IL-22 is a member of the IL-10 cytokine family, and can induce proliferative, antimicrobial and anti- apoptotic pathways on the intestinal epithelia, thus promoting tissue repair (Li et al., 2014). As such, this bispecific, termed 7A2-IgG4-IL22, may target and block inflammatory T cell homing and activation along the intestinal tract in areas of inflammation while promoting repair and protection from epithelial cells. In summary, 7A2-IgG4-IL22 represents one example of a new class of tissue-targeting, multimodal bispecifics to localize immunomodulating functions to inflamed tissues while minimizing systemic adverse effects.

## Results

### Design and Biochemical characterization of 7A2-IgG4-IL22

A gene encoding the bispecific, 7A2-IgG4-IL22, was engineered by fusing a previously described antagonistic anti-human MAdCAM-1 scFv 7A2 domain at the N-terminus of a human IgG4 Fc scaffold (Sparkes et al., 2025). The human IL-22 was introduced at the C-terminus of the IgG domain separated by a flexible GGGGSGGGGSGGGGSGGGGS linker sequence (Fig. 1A, left). IL-22 was selected as the therapeutic fusion partner in view of its reported role in promoting intestinal epithelial repair, barrier protection and antimicrobial peptide (AMP) production within the gastrointestinal tract (Li et al., 2014). The S228P mutation was inserted into the IgG4 Fc domain to prevent Fab arms exchange while preserving the structural stability of the IgG4 scaffold (Silva et al., 2015). The resulting construct was designed to cause its accumulation to inflamed areas of the intestinal tract by virtue of human MAdCAM-1 being principally expressed on the surface of endothelial cells located in the lamina propria. The bispecific was also engineered to display two therapeutic functions namely through its blockade of MAdCAM-1 binding to 4 7 integrin on T cells (anti-hMAdCAM- 1 7A2 scFv antagonistic domain) suppressing their activation and the promotion of beneficial epithelial responses through IL-22 signaling (human IL-22 domain).

**Figure 1.**
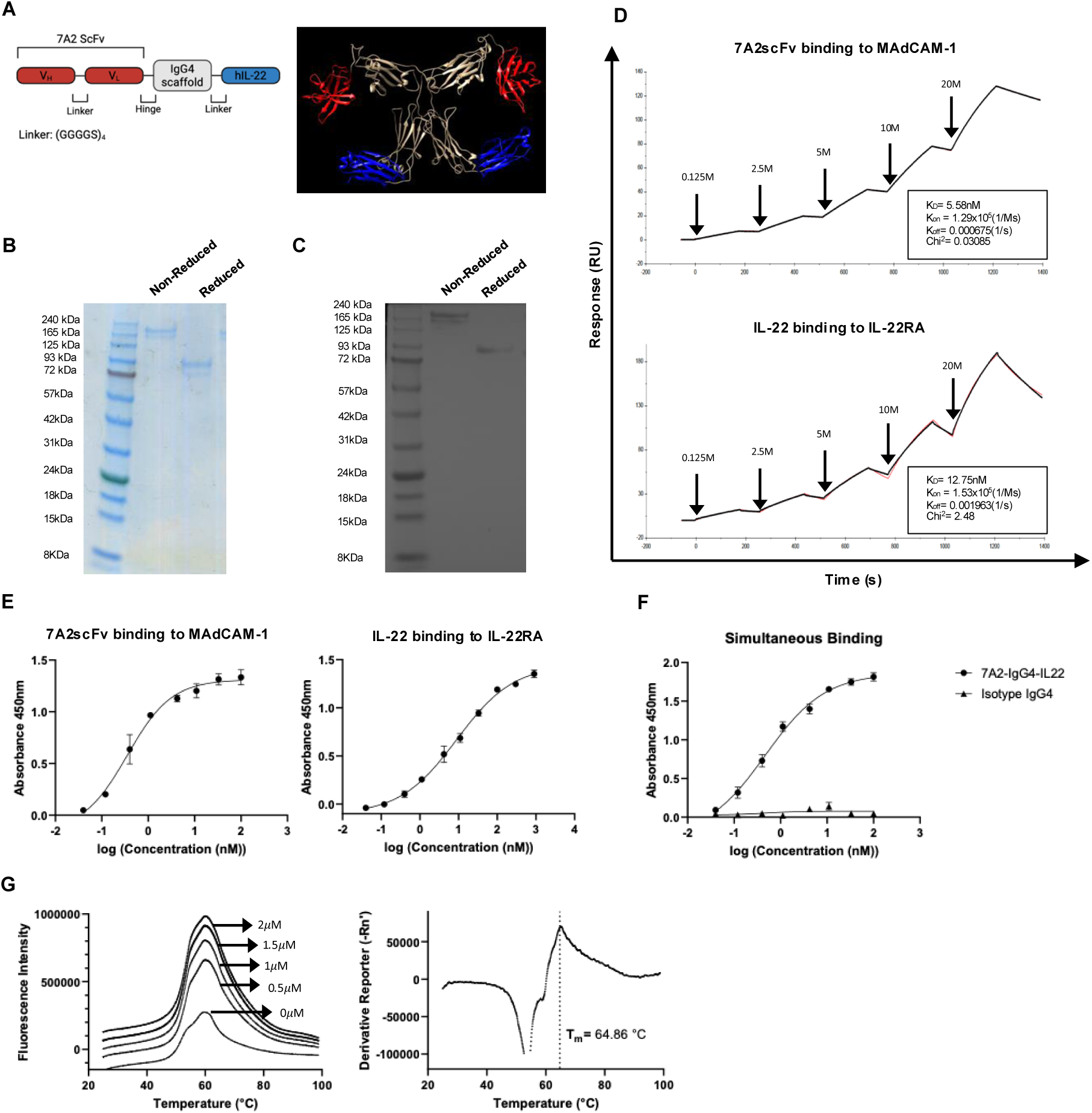
Biophysical and dual target binding characterization of 7A2-IgG4-IL22. **A)** Linear representation (left) and predicted ribbon (right) structure of 7A2-IgG4-IL22 bispecific as an Fc dimer using Chimera USCF software. **B)** Coomassie stained SDS-PAGE and **C)** Western blot of the purified bispecific under non-reduced and reduced conditions confirming the expected molecular weight and purity of the bispecific. **D)** Surface Plasmon Resonance (SPR) single-cycle kinetics sensorgrams (in red) and fitted curves (in black) with equilibrium (K_D_), association (k_on_), and dissociation (k_off_) rate constants depicting the binding of human MAdCAM-1 (top) and human IL-22RA (bottom) to 7A2-IgG4-IL22 immobilized onto a protein G sensor chip **E)** Concentration-dependent binding of the bispecific to individual targets. ELISA-based binding curves (450 nm) for anti human MAdCAM-1 7A2 scFv (left) and human IL-22 (right) binding to their respective immobilized targets. Data is presented as background-adjusted absorbance values fitted using a four-parameter logistic (4PL) nonlinear regression model. Each data point represents the average of technical triplicates +/- SD. **F)** Sandwich ELISA curve confirming the simultaneous dual-target binding of the bispecific. Wells were coated overnight with human IL22Rα1, and increasing amount of 7A2-IgG4-IL22 added to wells. Bound 7A2-IgG4-IL22 was detected using a biotinylated human MAdCAM-1 and HRP-streptavidin. Non- specific binding was measured using a human IgG4 isotype control. The measurements were performed in technical triplicates +/- SD. **G)** Thermal stability analysis of the bispecific using SYPRO orange. Left panel: background-adjusted fluorescence intensity as a function of temperature. Right panel: representative first derivative melt curve analysis used to determine the melting temperature (T_m_) at 1 *μ*M. Data is representative of 3 independent experiments performed in triplicates.

The bispecific was expressed as a secreted soluble protein in EXPI293F cells and purified by Protein G affinity chromatography followed by endotoxin removal. The predicted protein ribbon structure is depicted in Fig. 1A (right) using Chimera USCF software (Pettersen et al., 2004). Protein purity and integrity were confirmed by SDS-PAGE and Coomassie staining under both non-reducing and reducing conditions (Fig. 1B). Under non-reducing conditions, the construct migrated at approximately 170kDa, consistent with the expected molecular weight of the intact disulfide-linked bispecific molecule. Upon reduction, a predominant band was observed at approximately 93 kDa, corresponding to the dissociation of the interchain disulfide bonds and separation into monomeric chains. The SDS-PAGE elution profile in combination with a Western blot analysis of the 7A2-IgG4-IL22 bispecific using anti-human IgG heavy and light chain antibodies, confirmed the proper assembly and structural integrity of the engineered IgG4 Fc-based construct (Fig. 1C). The non-optimized expression of the bispecific by EXPI293F cells typically yielded ∼8-10 mg of the final, purified 7A2-IgG4-IL22 construct per liter of medium.

Binding kinetics were assessed by Surface Plasmon Resonance (SPR) analysis (Fig. 1D). The anti-MAdCAM-1 arm demonstrated high-affinity binding to human MAdCAM-1 with a dissociation constant (K_D_) of 5.6nM, an association rate constant (k_on_) of 1.29 x 10^5^ M^-1^ s^-1^, and a dissociation rate constant (k_off_) of 6.75 x 10^-4^ s^-1^. The observed K_D_ for the scFv 7A2 domain was partly reduced as compared to our previously reported full anti-MAdCAM-1 antibody 7A2 affinity (K_D_ = 0.68nM), consistent with a modest attenuation in binding resulting from re-engineering the F_ab_ chains into a single chain domain fused to a human IgG4 Fc scaffold (Fig. 1D, top) (Sparkes et al., 2025). 7A2-IgG4-IL22 bound to the human IL-22 receptor with a calculated K_D_ of 12.8nM (k_on_ = 1.53 x 10^5^ M^-1^ s^-1^ ; k_off_= 1.96x10^-3^ s^-1^), a value similar to the one previously reported for the IL-22-IL-22RA complex (20nM), indicating that the IL-22 binding to its receptor was preserved within our engineered 7A2-IgG4-IL22 construct (Fig. 1D, bottom) (Jones et al., 2008).

ELISA titrations were performed against immobilized human MAdCAM-1 and human IL-22RA to evaluate the binding of each domain to its respective target by the bispecific itself. The ELISA signal observed for the 7A2 scFv binding to human MAdCAM-1 plateaued around 10nM, consistent with the K_D_ reported by SPR (Fig. 1D, top; Fig. 1E, left). In contrast, binding to the IL-22 receptor saturated at higher concentrations, (>100nM), which is consistent with the comparatively higher k_off_ rate relative to MAdCAM-1 interacting with 7A2-IgG4-IL22 (Fig. 1E, right). To further assess simultaneous dual-target engagement, a bridging ELISA was performed using immobilized human IL-22 receptor. The binding of the bispecific was measured using biotinylated human MAdCAM-1 followed by Streptavidin-HRP (Fig. 1F). The bispecific demonstrated a clear dose-dependent increase in binding signal across the tested concentration range, with the ELISA signal saturating above 50nM. As expected, a baseline ELISA signal was observed for the IgG4 isotype control over the same concentration range (Fig. 1F). Collectively, these data demonstrate that the binding of each domain to their respective target within the engineered construct is preserved and can simultaneously bind to both MAdCAM-1 and the IL-22 receptor.

The thermal stability of the bispecific construct was also evaluated using the SYPRO Orange assay protein thermal shift assay. Fluorescence measurements as a function of temperature ranging from 20 °C to 99 °C were recorded over several protein concentrations to assess the unfolding behaviour and structural stability of 7A2-IgG4- IL22. Specifically, as temperature increased, 7A2-IgG4-IL22 progressively unfolded, exposing hydrophobic regions that bind SYPRO Orange dye, resulting in an increased fluorescence signal (thermal denaturation). Following 7A2-IgG4-IL22 fully unfolding, the fluorescence intensity declined at higher temperatures, consistent with aggregation-associated loss of accessible hydrophobic surfaces during protein denaturation (Elgert et al., 2020). Importantly, the unfolding transition remained constant across tested concentrations, indicating a structurally homogeneous and thermally stable protein with minimal evidence of conformational heterogeneity or aggregation prior to thermal denaturation (Fig. 1G, left). A first-derivative analysis of fluorescence change identified the melting temperature (T_m_) of 7A2-IgG4-IL22 to be 64.9 °C, corresponding to the point of maximal unfolding transition (Fig. 1G, right). The relatively high T_m_ value suggests that the human IgG4 Fc scaffold containing the S228P mutation, incorporating both the anti-MAdCAM-1 scFv 7A2 and human IL-22 domains, did not adversely affect protein stability or folding. Notably, this thermal stability is consistent with the range commonly reported for clinically developed IgG-based biologics, which frequently exhibit melting transitions between 60 and 70°C, supporting the biophysical robustness of the engineered construct (Jacobsen et al., 2017). Collectively, the preserved binding kinetics, robust dual-target binding, and favourable thermal stability profile of 7A2-IgG4- IL22 supported the further characterization of its functional properties.

### 7A2 scFv Blocks MAdCAM-1-mediated T-cell Activation, Proliferation, and Differentiation

To evaluate the functional consequences of MAdCAM-1 engagement on adaptive immune activation, primary human T cells were stimulated *in vitro* in the presence of plate-bound agonistic anti-CD3 monoclonal antibody (α -CD3 mAb) and recombinant human MAdCAM-1 and treated with the bispecific 7A2-IgG4-IL22 or an IgG4 isotype control (Fig. 2A). This assay was designed to better recapitulate intestinal-associated immune activation, where MAdCAM-1-mediated interactions contribute not only to lymphocyte trafficking but also to T cell co-stimulatory signaling within inflamed intestinal tissue (Girard et al., 2024; Lehnert et al., 1998).

**Figure 2.**
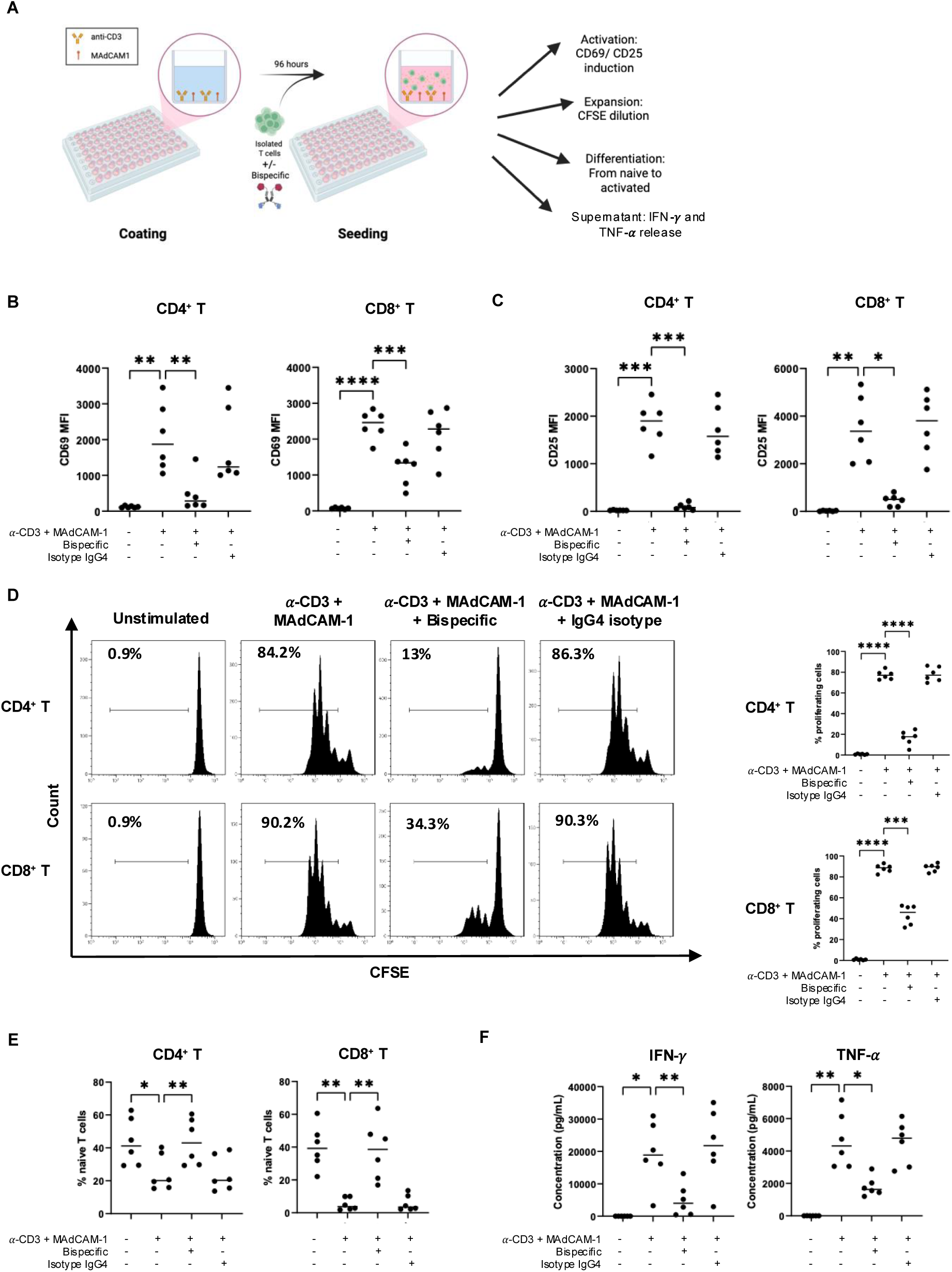
Functional characterization of the anti-MAdCAM-1 arm on T cell activation, proliferation, and phenotype. **A)** Schematic representation of the *in vitro* T cell activation assay. T cells were stimulated with anti-CD3 and MAdCAM- 1 +/- bispecific or IgG4 isotype control prior to downstream analysis. **B)** Expression of the early T cell activation marker CD69 on human CD4^+^ and CD8^+^ T cells, as determined by flow cytometry. Cells were stained 24h post stimulation. **C)** Expression of the late T cell activation marker CD25 on CD4**^+^** and CD8**^+^** T cells by flow cytometry. Cells were stained 96h post stimulation. **D)** Representative histograms of proliferating human CD4**^+^** and CD8**^+^**T cell percentages (left) and Quantification of T cell proliferation on CD4**^+^** and CD8**^+^** T cells based on CFSE cytometric dilution profiles (right) following a 96h post stimulation. **E)** Quantification of naïve CD4**^+^** and CD8**^+^** T cell frequencies based on CD45RA and CD27 expression. **F)** Quantification of IFN-*γ* and TNF-*α* release in supernatant of isolated T cells following a 96h post stimulation. *P-*values determined by repeated-measures one-way ANOVA, (\**P* <0.05, \*\**P* <0.005, \*\*\**P* <0.001). Results show the mean of 6 donors over 3 experiments, unless otherwise specified. MFI, median fluorescence intensity.

Mg^2+^ and retinoic acid were included in the culture system to better approximate intestinal-associated immune microenvironmental conditions. Specifically, magnesium supports integrin-dependent adhesion and signalling, whereas retinoic acid is a key intestinal-associated metabolite involved in T cell trafficking by inducing α 4β7 integrin and CCR9 expression (Girard et al., 2024; Su et al., 2023). Flow cytometry was performed to identify and characterize human CD4^+^ and CD8^+^ T cell populations upon stimulation by the MAdCAM-1 axis, which leads to their activation, expansion and differentiation (Suppl. Fig. 1A).

CD69 and CD25 were selected as complementary activation markers based on their distinct activation kinetics. CD69 is rapidly upregulated within hours on both CD4^+^ and CD8^+^ T cell populations following T cell receptor engagement of isolated human T cells with α -CD3 mAb and MAdCAM-1, reflecting early proximal activation signaling (Fig. 2B). MAdCAM-1 co-stimulation enhanced CD69 expression in both T cell subsets in comparison to unstimulated controls, consistent with early activation signaling. Treatment with the bispecific significantly reduced CD69 expression compared to the IgG4 isotype control on both T cell subsets, indicating effective blockade of MAdCAM-1- mediated activation (Fig. 2B; 24-hour stimulation). In contrast, CD25 expression increases over 24-72 hours and is associated with sustained IL-2 signaling, proliferative commitment, and progression towards effector differentiation (Craston et al., 1997; Motamedi et al., 2016). As such, late activation was evaluated by CD25 expression on both T cell subsets after 4-day stimulation. MAdCAM-1 stimulation increased CD25 expression in both CD4^+^ and CD8^+^ T cells, reflecting enhanced IL-2 signaling and sustained activation. Bispecific treatment reduced CD25 expression relative to IgG4 isotype control in both T cell subsets, indicating the attenuation of prolonged activation signaling (Fig. 2C).

Next, T cell proliferation was measured by flow cytometry using CFSE dilution, with a view to assess if 7A2-IgG4-IL22 can attenuate the polyclonal expansion of potentially inflammatory T cells. Cell proliferation usually occurs after 48-72 hours and was assessed in the present study after 4 days of stimulation (Li et al., 2007; Marinescu et al., 2021). As expected, α -CD3 mAb + MAdCAM-1 stimulation enhanced the proliferative expansion of both CD4^+^ and CD8^+^ T cells relative to unstimulated controls, while adding the bispecific significantly reduced proliferation when compared to the IgG4 control (Fig. 3D, left). Quantification of proliferating human T cells confirmed a remarkable reduction of both T cell subset frequencies from 77% to 17% for CD4^+^ T cells and from 88& to 44% for CD8^+^ T cells following their treatment with the bispecific (Fig. 3D, right).

**Figure 3.**
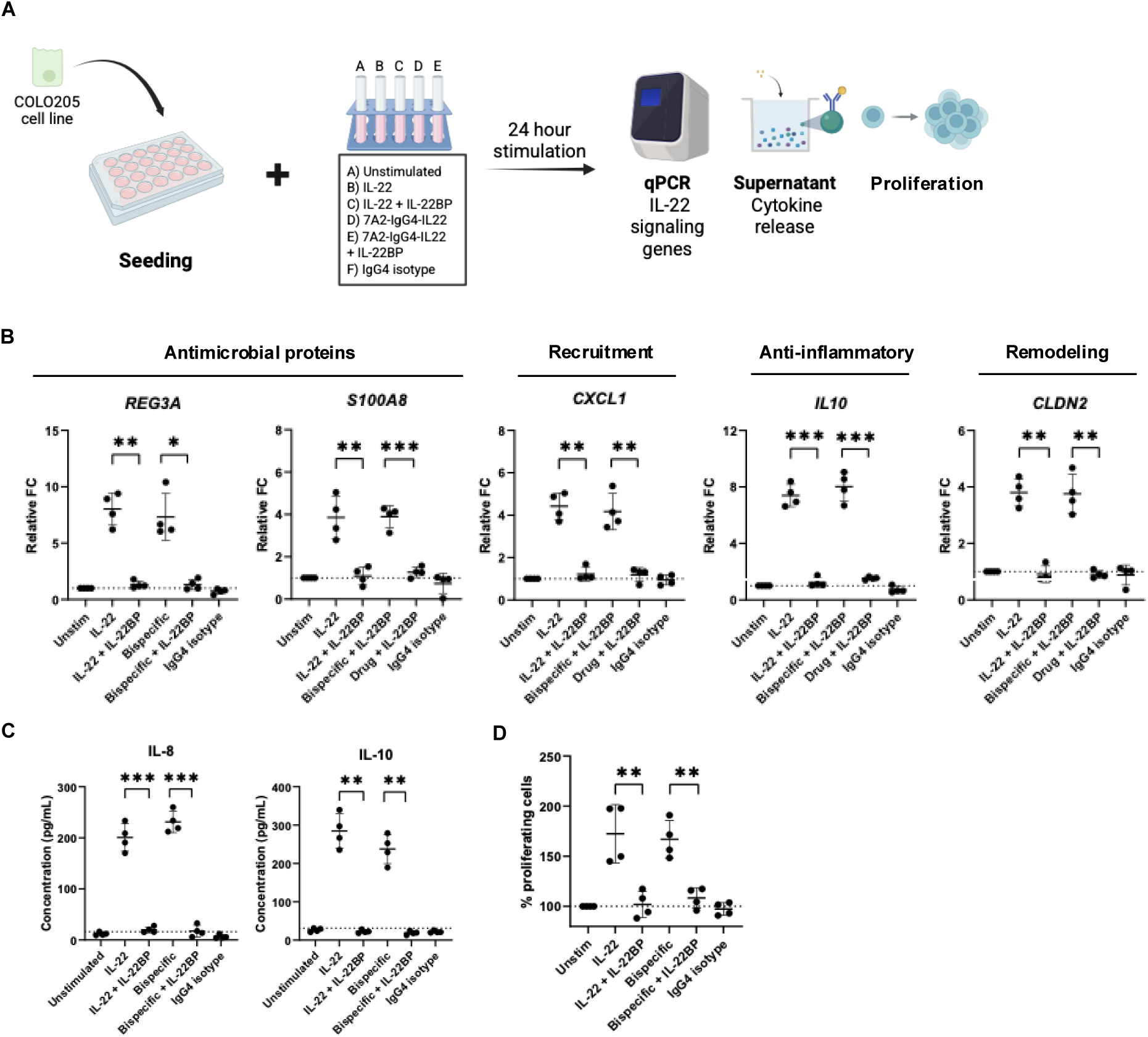
7A2-IgG4-IL-22 induces IL-22 response on COLO205 cells *in vitro*. **A)** Schematic of protocol for 24-hour COLO205 cell stimulation with IL-22/7A2-IgG4-IL-22 +/- IL-22BP. **B)** Real-time qPCR analyses of relative fold change in expression of selected mRNA genes by stimulated COLO205 cells using IL-22 +/- IL-22BP signaling. **C)** Cytokines released by COLO205 cells upon IL-22 signaling +/- IL-22BP. **D)** MTT assay of stimulated COLO205 cells showing IL-22 mediated epithelial metabolic and proliferating responses. Data is presented as mean +/- SD. Results showed are representative of **n=4** experiments, unless otherwise specified. *P-*values determined by paired t test, (\**P* <0.05, \*\**P* <0.005, \*\*\**P* <0.001).

The phenotypic differentiation of these T cell subsets was subsequently assessed based on CD45RA and CD27 expression. Naïve T cells were defined as CD45^+^CD27^+^; central memory-like cells as CD45^-^CD27^+^; effector memory-like cells as CD45RA^-^CD27^-^; and terminally differentiated cells as CD45RA^+^CD27^-^ (Suppl. Fig 1B) (Muroyama and Wherry, 2021). Under unstimulated conditions, both CD4^+^ and CD8^+^ T cell compartments remained predominantly naïve (Fig. 2E). Anti-CD3 + MAdCAM-1 stimulation reduced the proportion of naïve T cells (CD45^+^CD27^+^), consistent with activation-driven differentiation. Treatment with the bispecific antibody significantly preserved the naïve T cell compartment relative to IgG4 control (Fig. 2E), suggesting a strong attenuation of the activation-to-differentiation transition.

To determine whether this activation program translates into effector cytokine production, supernatants collected after 4 days of stimulation were analyzed using multiplex bead-based cytokine quantification. Anti-CD3 + MAdCAM-1 stimulation of human T cells increased the secretion of key inflammatory cytokines IFN and TNF as compared to unstimulated conditions. Importantly, treatment with the bispecific reduced the levels of both cytokines relative to IgG4 control (Fig. 3F). Given that IFN and TNF are central mediators of epithelial damage and chronic inflammation in IBD, these findings indicate that MAdCAM-1 co-stimulation enhances a pathogenic effector cytokine program that is effectively attenuated by the bispecific (Funderburg et al., 2013; Imam et al., 2018; Souza et al., 2023). Collectively, these data demonstrates that MAdCAM-1 co-stimulation acts in concert with CD3 signaling to enhance early activation, sustain IL-2-dependent signaling, promote time-dependent proliferation, drive differentiation away from a naïve phenotype, and induce secretion of key inflammatory cytokines (Girard et al., 2024; Lehnert et al., 1998; Vimonpatranon et al., 2023). Blockade of MAdCAM-1 signaling using the bispecific attenuates all of these processes, supporting a central role for MAdCAM-1 in amplifying pathogenic T cell activation programs relevant to intestinal inflammation.

### The IL-22 arm of the bispecific 7A2-IgG4-IL22 induces epithelial repair-associated signaling and antimicrobial transcriptional programs

To evaluate epithelial responses mediated by the IL-22 axis, human COLO205 intestinal epithelial cells were treated with recombinant human IL-22 alone, human IL-22 in the presence of human IL-22BP, the bispecific 7A2-IgG4-IL22, the bispecific in the presence of human IL-22BP, or an IgG4 isotype control to assess cytokine-driven repair and inflammatory signaling (Fig. 3A). IL-22BP is a soluble endogenous antagonist that binds to IL-22 tightly and prevents its interaction with the IL-22 receptor complex, thereby impairing downstream IL-22-mediated signalling pathways (Zenewicz, 2021). COLO205 cells were selected because of their high responsiveness to IL-22-mediated epithelial signaling and suitability for evaluating cytokine-driven transcriptional and metabolic responses (Arshad et al., 2020; Nagalakshmi et al., 2004).

To define downstream epithelial transcriptional programs being involved, qPCR analysis was performed to assess gene expression of IL-22-induced signaling such as antimicrobial defense, barrier function and epithelial immune signaling (Zenewicz, 2018a). IL-22 stimulation upregulated key antimicrobial and epithelial defense genes, including *REG3A* and *S100A8*, consistent with activation of mucosal protective responses (Fig. 3B) (Faivre et al., 2025; Hendrikx et al., 2020; Zenewicz, 2018a). In parallel, IL-22 also modulated genes associated with immune cell recruitment, communication and epithelial remodeling, including *CXCL1, IL10* and *CLDN2* (Fig. 3B) (Nagalakshmi et al., 2004; Pavlidis et al., 2022; Wang et al., 2017a; Young et al., 2017). Notably, similar transcriptional responses were observed following treatment with 7A2- IgG4-IL22, supporting the preservation of functional IL-22 signaling within the engineered therapeutic format. Co-treatment with IL-22BP attenuated these responses, confirming dependence on IL-22 receptor engagement (Fig. 3B)

In addition, changes in epithelial cytokine secretion were assessed following cytokine stimulation. Both recombinant human IL-22 and the bispecific induced release of IL-8 and IL-10, whereas this response was completely attenuated in the presence of IL-22BP (Fig. 3C). The simultaneous induction of a proinflammatory cytokine such as IL- 8 together with the regulatory cytokine, IL-10, is consistent with the pleiotropic nature of IL-22 signaling within epithelial tissues (Dudakov et al., 2015; Keir et al., 2020). While IL-22 promotes antimicrobial defense and recruitment of immune cells that may be involved in tissue repair and mucosal protection, it can also activate compensatory regulatory pathways that help limit excessive inflammation and support restoration of intestinal homeostasis.

Finally, COLO205 epithelial cells metabolic activity was assessed using an MTT assay, in which mitochondrial oxidoreductase enzymes reduce the tetrazolium salt MTT into insoluble formazan crystals. The amount of formazan produced is proportional to the metabolic activity of viable cells and is commonly used as an indirect indicator of cell viability and proliferation, as an increase in cell number is typically accompanied by an increase in total cellular metabolic activity. This assay was used to confirm that IL-22 signaling did lead to downstream proliferative and regenerative responses in these cells (Senthilraja and Kathiresan, 2015). IL-22 has previously been reported to promote such functions following mucosal injury (Keir et al., 2020; Li et al., 2014). Restoration of epithelial integrity is particularly relevant in IBD, where disruption of the intestinal barrier and development of a “leaky gut” state are key pathological features that contribute to sustained inflammation and microbial translocation (Imam et al., 2018; Yue et al., 2024). Both recombinant IL-22 and 7A2-IgG4-IL22 increased epithelial metabolic activity compared to unstimulated or IgG4 isotype control, whereas this response was attenuated in the presence of IL22BP, further supporting the preservation of functional IL-22 signaling within the engineered construct (Fig. 3D). Collectively, these findings demonstrate that the human IL-22 arm of the bispecific 7A2-IgG4-IL22 remains biologically functional and recapitulates canonical IL-22 epithelial responses, including antimicrobial defense gene induction, epithelial immune signaling, and enhanced epithelial metabolic activity, supporting a potential role for the bispecific in promoting mucosal repair pathways relevant to barrier restoration.

## Discussion

Inflammatory bowel disease (IBD) affects approximately 3.8 million people worldwide and places patients at increased risk of severe long-term complications, including colorectal cancer (CRC), with immune dysregulation playing a key role in disease progression as well as chronic mucosal injury (Ruan et al., 2025). Although several anti-integrin therapeutic approaches have demonstrated strong clinical efficacy in IBD, limitations associated with systemic distribution and off-target immunosuppression represent significant concerns (Kelly and Long, 2024;Chu et al., 2023; D’Haens et al., 2018; Wang et al., 2018; Yeshi et al., 2024). Broad inhibition of leukocyte trafficking has been associated with severe adverse effects, including increased susceptibility to opportunistic infections such as progressive multifocal leukoencephalopathy (PML) (Domènech and Gisbert, 2016; Vermeire et al., 2024; Yeshi et al., 2024). These limitations highlight the need for next-generation therapeutic strategies capable of providing localized immune modulation while simultaneously promoting epithelial restoration within the intestinal microenvironment.

In the present study, we developed and characterized a gut-targeted IgG4-based bispecific termed 7A2-IgG4-IL22 that combines an anti-MAdCAM-1 targeting arm with a human IL-22 cargo. Rather than functioning solely as a tissue-localizing arm, our findings support a broader role for MAdCAM-1 as a co-stimulatory amplifier of pathogenic T cell activation. Specifically, co-stimulation through CD3 and MAdCAM-1 has been shown to promote early and late activation marker expression, enhanced T cell expansion, and drove differentiation away from naïve state (Fig. 2B, 2C, 2D and 2E) (Girard et al., 2024; Grant et al., 2001; Vimonpatranon et al., 2023). This activation program led to the enhanced secretion of IFN and TNF, two cytokines strongly implicated in epithelial barrier dysfunction and chronic intestinal inflammation (Funderburg et al., 2013; Imam et al., 2018; Souza et al., 2023).

Current therapies such as adalimumab (Humira) primarily neutralize circulating TNF systemically and have substantially improved the clinical management of IBD. However, broad systemic suppression of TNF can impair physiological immune surveillance and host defense mechanisms, increasing susceptibility to opportunistic infections and other immune-related adverse effects (Assche et al., 2007; Chao and Liao, 2025; Vural et al., 2023). In addition, systemic cytokine blockade does not selectively target inflamed intestinal tissue and may suppress immune activity in non-disease tissues. In contrast, our bispecific limits activation of tissue-associated T cell via the MAdCAM-1 axis, thus simultaneously reducing both IFN and TNF production in a localized manner. This distinction is biologically important, as TNF and IFN synergistically promote epithelial apoptosis, tight junction disruption and sustained inflammatory signaling within the intestinal mucosa (Woznicki et al., 2021). Reduction of these cytokines is therefore expected to alleviate epithelial stress, reduce barrier permeability, and interrupt inflammatory amplification loops that perpetuate chronic disease while potentially minimizing systemic immune suppression. Notably, indirect dampening of mucosal inflammatory cytokine programs has also been reported following the blockade of the MAdCAM-1/ 4 7 integrin axis with ontamalimab and vedolizumab, where reduced recruitment of inflammatory lymphocytes into lamina propria was associated with decreased expression of T cell activation-related gene signatures. These findings suggest that localized inhibition of intestinal trafficking can attenuate inflammatory cytokine networks without the need for systemic cytokine neutralization, representing a potential advantage over anti-TNF therapies that broadly suppresses circulating cytokines. While these effects are thought to arise primarily from reduced immune cell infiltration, our bispecific additionally demonstrated direct inhibition of MAdCAM-1-mediated T cell activation together with localized IL-22 delivery, thereby integrating immune modulation with epithelial restoration (Schulze et al., 2023). Furthermore, attenuation of Th1-associated cytokines may indirectly reduce the downstream activation of innate immune pathways, including inflammasome-associated cytokines such as IL-18 and IL-1, which contribute to myeloid cell-driven propagation of inflammatory responses. Both cytokines have been implicated in epithelial barrier dysfunction and are known to modulate adaptive immune responses, including Th17 differentiation and maintenance of inflammatory T cell programs within the intestinal microenvironment. Importantly, production of IL-18 and IL-1 can be triggered by gut dysbiosis and microbial translocation resulting from epithelial barrier disruption, thereby reinforcing chronic inflammatory signaling loops in IBD (Aggeletopoulou et al., 2024; Mokry et al., 2019; Nowarski et al., 2015). In this context, localized intestinal targeting through the MAdCAM-1 axis may provide an opportunity not only to suppress pathogenic T cell activation and trafficking, but also to modulate innate immune compartments involved in sustaining mucosal inflammation (Briskin et al., 1997; Girard et al., 2024; Grant et al., 2001; Lehnert et al., 1998; Tan et al., 1998; Vimonpatranon et al., 2023). Future adaptations of this modular platform could therefore incorporate cargos specifically designed to target myeloid-associated inflammatory pathways within the inflamed intestine.

Beyond immune suppression, a major advantage of the present bispecific lies in its incorporation of human IL-22, a cytokine known to mediate epithelial repair signaling. IL-22 has been widely reported to promote epithelial regeneration, antimicrobial peptide (AMP) production, and mucosal barrier protection (Li et al., 2014). Consistent with this biology, the IL-22 arm of the bispecific induced transcriptional programs associated with epithelial defense and repair, including *REG3A, S100A8, CLDN2, CXCL1* and *IL10* (Zenewicz, 2018b). Upregulation of Reg3 family members and calgranulin-A (*S100A8*) support enhanced AMP production, an important component of mucosal defense against microbial invasion, especially in IBD, where the epithelial barrier is compromised (Faivre et al., 2025; Hendrikx et al., 2020; Zenewicz, 2018b). Induction of Claudin 2 (*CLDN2*) may additionally reflect active epithelial remodeling and restitution processes, as Claudin-2 has been implicated in regulation of epithelial turnover, wound healing, and controlled restoration of mucosal barrier dynamics during intestinal injury responses in IBD (Wang et al., 2017b). In particular, IL-10 induction favors the activation of compensatory anti-inflammatory epithelial programs, while the expression of *CXCL1* likely reflects controlled chemokine-mediated recruitment involved in tissue surveillance and repair (Nagalakshmi et al., 2004; Pavlidis et al., 2022; Young et al., 2017). Together, these findings support the concept that IL-22 signaling induces a coordinated epithelial response integrating antimicrobial defense, barrier restoration and regulated immune response.

Another important design component of 7A2-IgG4-IL22 is the use of a human IgG Fc scaffold, which is expected to improve the pharmacokinetic profile through FcRn-mediated recycling, thus increasing its half-life and exposure compared to an Fc-free format (Czajkowsky et al., 2012). Unlike IgG1-based therapeutic antibodies, the human IgG4 Fc domain reduces Fc-mediated effector function resulting in lower complement activation and diminished engagement of Fc receptors associated with antibody-dependent cellular cytotoxicity and inflammatory immune activation (Piseddu et al., 2025). This distinction may be particularly important in IBD, where macrophages and myeloid-populations play central roles in coordinating intestinal inflammation through cytokine production and antigen presentation (Bain Allan McI Mowat et al., 2014; Flannigan et al., 2015). Reduced Fc receptor engagement may therefore limit unintended macrophage activation and downstream inflammatory amplification within the intestinal mucosa (Castro-Dopico and Clatworthy, 2019). Consequently, this human IgG4 Fc domain is well suited for therapeutic strategies aimed at localized immunomodulation rather than cell depletion approaches. Additionally, the MAdCAM-1 targeting component within 7A2-IgG4-IL22 may allow for the preferential localization of the construct to inflamed intestinal vasculature, potentially reducing systemic exposure and limiting off-target immunosuppression. However, the use of an IgG4 scaffold also presents important limitations that should be considered. Reduced Fc-mediated effector function, while advantageous for minimizing inflammatory activation, may also alter immune complex clearance compared to IgG1-based therapeutics. Furthermore, we incorporated a stabilizing S288P mutation within the human IgG4 Fc domain to reduce Fab arms exchange. Structural heterogeneity and long-term stability remain important considerations during therapeutic development and large-scale manufacturing (Silva et al., 2015). Additional studies evaluating pharmacokinetics, tissue retention, immunogenicity, and long-term safety will therefore be necessary to further assess the translational suitability of this platform *in vivo*.

Importantly, the modular architecture of this platform suggests future development of “plug-and-play” gut targeted therapeutics. While IL-22 was selected here because of its established role in epithelial protection and regeneration, alternative cargos such as IL-10 and IL-18BP could theoretically be incorporated to modulate distinct inflammatory pathways or immune compartments (Li et al., 2014). Future iterations of this platform could potentially target innate immune populations, including monocytes and macrophages, which orchestrate inflammation through cytokines such as IL-12, IL-1 and IL-18 (Bain Allan McI Mowat et al., 2014; Maerten et al., 2004). Moreover, the versality of this strategy may extend beyond IBD towards other intestinal diseases, including inflammation-driven colorectal cancer, where chronic immune activation and epithelial remodeling coexist.

Finally, restoration of epithelial barrier integrity may have broader implications beyond local tissue repair. Reduction of epithelial permeability could decrease microbial translocation and exposure to luminal pathogen-associated molecular patterns, thereby limiting secondary activation of innate immunity that sustain chronic inflammation (Mukherjee, 2026). Collectively, these results support a therapeutic strategy that integrates localized immunosuppression with epithelial repair and protection and establish proof-of-concept for a modular gut-targeted bispecific platform in IBD.

## Methods

### Sex as a biological variable

Sex was not considered as a biological variable.

### Statistical analysis

All data is presented as the result of four to six independent experiments and expressed as mean +/- SD. GraphPad Prism v.9.3.1 (GraphPad Software, San Diego, CA, USA). The statistical analysis performed for each experiment is included in the figure legend. P <0.05 was defined as statistically significant. Outlier analysis was performed using the ROUT method (Q = 1%). Number of replicates are indicated in the respective results sections and figure legends.

## Authors contributions

JDS and AS designed experimental methods. JDS, NA, and JCL performed technical work. JDS performed experiments and data analysis. AS and JCL enrolled patients for the study. JDS and JG wrote the main manuscript text and prepared the figures. All authors reviewed the manuscript.

## Data Availability

All data produced in the present study are available upon reasonable request to the authors

## Acknowledgments

We would like to recognize the support of the healthcare team at the Sunnybrook Health Science Centre Transfusion Medicine Clinic for their assistance with sample collection, as well as all blood donors whose participation made this study possible. We would also like to recognize Dr. Rima Al-Awar for generously providing the COLO205 cell line.

## Funding Sources

This work was supported by the Canadian Institutes of Health Research (CIHR) project grants PJT-468907 and PJT-517882 to JG.

---

**Suppl. Figure 1.**
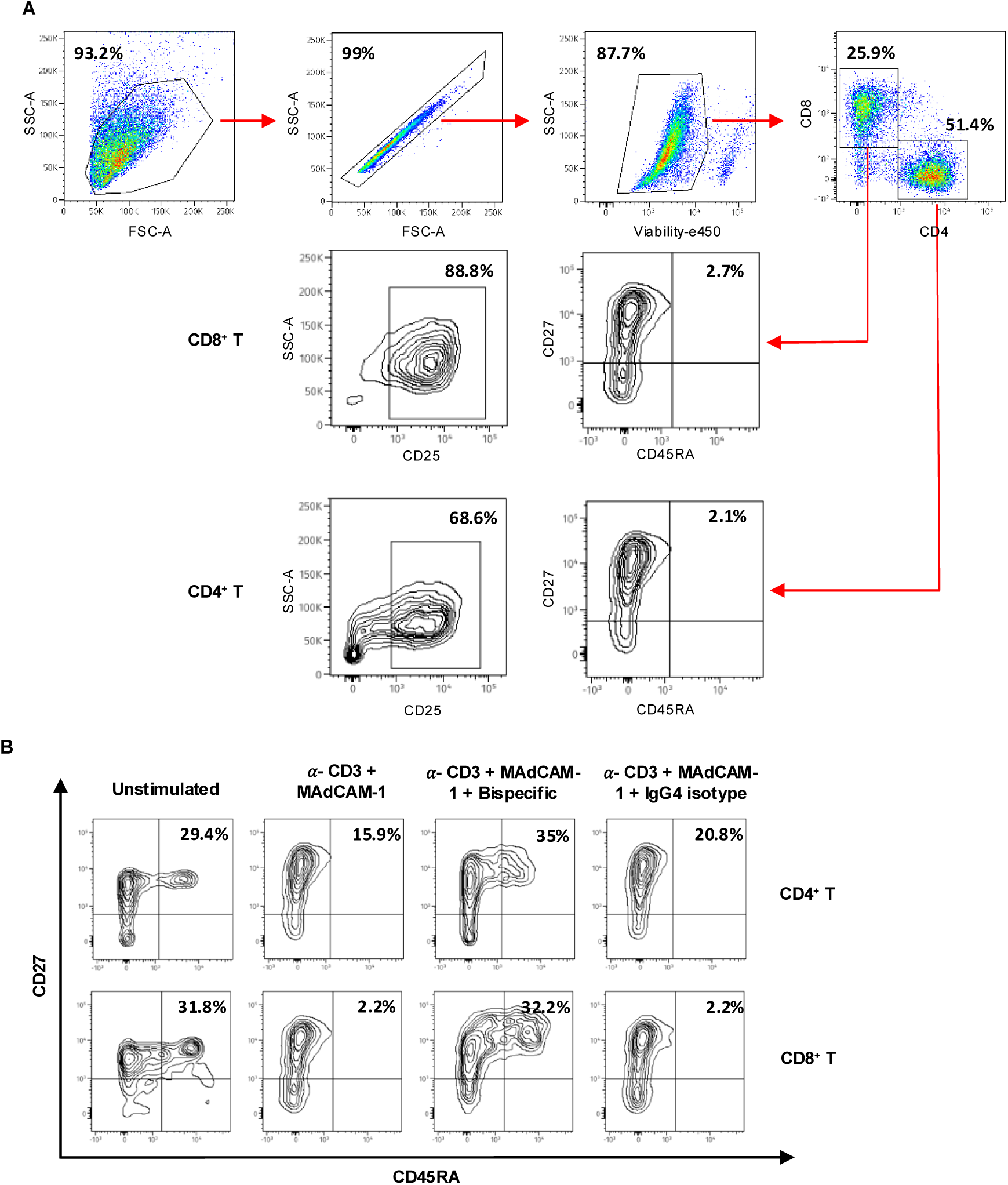
Flow cytometry gating and phenotypic characterization of T cell activation assay. **A)** Representative flow cytometry gating strategy used to identify CD4^+^ and CD8^+^ T cell populations for downstream activation, and differentiation analyses. **B)** Representative contour plots showing T cell differentiation states based on CD45RA and CD27 expression. Naïve T cells were defined as CD45RA^+^CD27^+^; central memory-like cells as CD45RA^-^CD27^+^; effector memory-like cells as CD45RA^-^CD27^-^; and terminally differentiated cells as CD45RA^+^CD27^-^.

## Notes

### Competing Interest Statement

The authors have declared no competing interest.

### Author Declarations

Informed consent was obtained from all subjects. All collections were performed in accordance with the guidelines and regulations set by the Institutional Sunnybrook Research Ethics Board under Research Ethics Board approval 2978.

