## Supplementary information for "A Gut-Specific Bispecific Combining MAdCAM-1 Blockade and IL-22 Signaling to Halt T-Cell Inflammation and Promote Mucosal Restoration"

**Lead contact**

Further information and requests for resources and reagents should be directed to, and will be fulfilled by, the Lead Contact, Jean Gariépy

**Human donors and peripheral blood mononuclear cells (PBMCs) collection**

Six healthy human donors were included to obtain PBMCs from whole blood. Informed consent was obtained from all subjects. All collections were performed in accordance with the guidelines and regulations set by the Institutional Sunnybrook Research Ethics Board under Research Ethics Board approval #2978.

**Methods details**

MAdCAM-1 targeting bispecific generation

A gene encoding the human MAdCAM-1 targeting 7A2-IgG4-IL22 bispecific was generated where the resulting protein has a scFv derived from the blocking anti-human MAdCAM-1 monoclonal antibody 7A2 fused to the N-terminus of a human IgG4 Fc domain followed by a human IL-22 (Q9GZX6-1) at the C-terminus of the IgG domain using a flexible GGGGSGGGGSGGGGSGGGGS linker sequence, as depicted in Figure 1A. The gene was cloned into a pcDNA3.4 TOPO TA mammalian expression plasmid (Thermo Fisher Scientific) and expressed in Expi293F cells (Thermo Fisher Scientific, Cat: A14527) using the Expi-293 mammalian cell expression system (Thermo Fisher Scientific, Cat: A1435101). A 5′ Ig-κ leader sequence was included in the construct for high protein secretion by mammalian cells. The secreted protein was subsequently purified by affinity chromatography using a HiTrap protein G high performance column (Cytiva, Cat: 29-0485-81). 7A2-IgG4-IL22 was eluted with 0.1M glycine HCl pH 2.7 and desalted into PBS using PD-10 columns (Cytiva, Cat: 45000148).

Endotoxin removal

Following purification, endotoxin removal was performed using Pierce^TM^ High-capacity endotoxin removal spin columns (Thermo Fisher Scientific, Cat: 88277) according to manufacturer’s batch incubation protocol. Briefly, the resin was equilibrated prior to use with 0.2N NaOH overnight at room temperature. This step was followed by sequential washes with 2M NaCl, endotoxin-free ultra-pure water, and endotoxin-free phosphate-buffered saline (PBS) (Wisent, Cat: 311-011-CL). Purified 7A2-IgG4-IL22 was added directly to the equilibrated resin and incubated 1 hour at 4°C with gentle end-over-end mixing to facilitate endotoxin binding. The column was subsequently centrifuged at 500 x g for 1 minute to recover the endotoxin-depleted 7A2-IgG4-IL22 and its concentration was quantified using a NanoDrop 2000 (ThermoFisher).

Protein modelling

A predicted ribbon structure of 7A2-IgG4-IL22 was generated using UCSF Chimera software, developed by the resource for Biocomputing, Visualization, and Informatics at the University of California, San Francisco, with support from NIH P41-GM103311. (Pettersen, et al., 2004).

Biochemical characterization of the bispecific 7A2-IgG4-IL22

The apparent molecular weight, dimeric state and purity of the bispecific, 7A2-IgG4-IL22 were confirmed by SDS-PAGE by loading 0.5µg of the bispecific onto a Bolt 4-12%, Bis-Tris Plus gel (Invitrogen, Cat: NW04120BOX). The protein samples were electrophoresed at 200V for 35 minutes under non-reducing and reducing conditions. Following electrophoresis, the gel was vertically divided into two halves, each containing BLUelf prestained protein ladder (FroggaBio, Cat: PM008-0500F). One half of the gel was stained with InstantBlue Coomassie protein stain (abcam, Cat: 50-196-3787) to assess protein purity and molecular weight, while the other half was transferred onto an Amersham^TM^Protran^TM^ 0.45-μm nitrocellulose blotting membrane (Cytiva, Cat: 10600002) using transfer buffer containing 48 mM Tris base, 39 mM glycine, 20 % v/v methanol. The membrane was blocked overnight in 5% milk at 4°C and subsequently incubated with goat anti-human IgG Fc fragment antibody conjugated to HRP (Bethyl, Cat: A80-104P) diluted at 1:10,000 at room temperature for 1 hour. The membrane was washed with Tris Buffer Saline (TBS) + 0.1% Tween 20 (TBST) prior to development using Pierce^TM^ 1-step Ultra TMB blotting solution (Thermo Fisher Scientific, Cat: 37574). The blot was imaged using a Fusion FX chemiluminescent imager (Vilber) with a 40-milisecond exposure.

Surface plasmon resonance

Surface Plasmon Resonance (SPR) was used to determine the separate binding kinetics of 7A2-IgG4-IL22 to full-length recombinant human MAdCAM-1 extracellular domain (Sparkes, et al., 2025) and human IL-22RA. SPR experiments were performed on a BiaCore T200 (Cytiva) using a protein G sensor chip (Cytiva, Cat: 29179315). 7A2-IgG4-IL22 was immobilized on the chip via its Fc domain at a concentration of 20µg/mL. Kinetic measurements were performed using single-cycle kinetics with five concentrations of human MAdCAM-1 generated by 1:2 serial dilutions starting at 20nM. Analyte injections were performed at a flow rate of 30µL/min for 300s per concentration, followed by an association phase and dissociation phase. RU values were additionally recorded for 180s followed by regeneration. The system was run in HBS-EP running buffer (20mM HEPES pH 7.4, 150mM NaCl, 0.005% Tween-20, 3.4mM EDTA). Binding parameters were also derived from SPR sensorgrams for human IL22RA interacting with 7A2-IgG4-IL22 immobilized of the same protein G sensor chip. Sensor surfaces were regenerated between cycles using a regeneration solution (10nM Glycine-HCl, pH 1.7 solution) (Cytiva, Cat: BR100354) and reference sensorgram responses were subtracted prior to analysis. The sensorgrams were fitted using a 1:1 Langmuir binding model to determine the equilibrium dissociation constant (K_D_), association rate constant (K_on_), and dissociation rate constant (K_off_).

ELISA assays

Concentration-dependent curves were generated by ELISA to evaluate the binding of 7A2-IgG4-IL22 to human MAdCAM-1 and human IL22Rα. For the anti-MAdCAM-1 binding assay, 1μg/ml of human MAdCAM-1 was coated onto high-bind ELISA plate (Corning, Cat: 07200642) and incubated overnight at 4°C (Sparkes, et al., 2025). For the IL-22 binding assay, 1μg/ml of recombinant human IL22Rα1 (R&D Systems, Cat: 2770-LR-050) was coated under the same conditions. The plate was blocked with 1% BSA at room temperature for 1hr.

Serial dilutions of 7A2-IgG4-IL22 (1:3) were added to individual wells starting at 100nM and the plate was incubated for one hour at room temperature. A human IgG4 isotype control (BioLegend, Cat: 403701) was used to assess any non-specific binding. Following incubation, bound 7A2-IgG4-IL22 was detected with a goat anti-human IgG Fc fragment antibody conjugated to HRP (1:10,000 dilution; Bethyl, Cat: A80-104P) and incubated for 45 minutes at room temperature. The plate was washed 3 times between each step with PBST (PBS + 0.05% Tween 20). One-step^TM^ Ultra TMB-ELISA (Thermo Fisher Scientific, Cat: 34028) was dispensed into each well and neutralized with 1:3 dilution of 0.16M sulfuric acid. Absorbance was measured at 450 nm using a Synergy H1 microplate reader. Optical density values were blank-adjusted prior to analysis and dose-response curves were fitted using a four-parameter logistic (4PL) sigmoidal model to determine saturation/ plateau binding concentrations. Average optical density measurements ± SD were derived from experiments performed in triplicate.

Sandwich ELISA

A sandwich ELISA was performed to determine if 7A2-IgG4-IL22 can simultaneously bind human IL22Rα and human MAdCAM-1. Recombinant human IL22Rα1 (1μg/ml; R&D Systems, Cat: 2770-LR-050) was dispensed into wells of an ELISA plate (Corning, Cat: 07200642) and incubated overnight at 4°C. The plate was blocked with 1% BSA at room temperature for 1hr. The bispecific 7A2-IgG4-IL22 was added to wells covering a range of 9 concentrations (1:3 dilution) starting at 100nM and the plate was incubated for one hour at room temperature. Non-specific binding was measured using a human IgG4 isotype control (BioLegend, Cat: 403701) was used to assess. Following this incubation step, biotinylated human MAdCAM-1 (1μg/ml) was dispensed into each well and allowed to bind for one hour at room temperature. Finally, an avidin-HRP conjugate (1:1,000 dilution; BioLegend, Cat: 405103) was added to wells and incubated for 45 minutes at room temperature. The plate was washed 3 times between each step with PBST (PBS + 0.05% Tween20). One-step^TM^ Ultra TMB-ELISA (Thermo Fisher Scientific, Cat: 34028) was dispensed into each well for one minute and the reaction was neutralized with 1:3 dilution of sulfuric acid. Absorbance was measured at 450 nm using a Synergy H1 microplate reader. Optical density values were blank-adjusted prior to analysis. Dose-response curves were fitted using a four-parameter logistic (4PL) sigmoidal model to determine saturation/ plateau binding concentrations. Average optical density measurements ± SD were derived from experiments performed in triplicate.

Thermal shift assay

A thermal shift assay (TSA) was performed to evaluate the thermal stability of 7A2-IgG4-IL22 using SYPRO™ Orange protein gel stain (Invitrogen, Cat: S6650). Reactions containing 7A2-IgG4-IL22 at final concentrations of 2.5, 2, 1.5, 1, 0.5 μM as well as in the absence of the bispecific, were prepared in the presence of SYPRO™ Orange protein gel stain according to the manufacturer’s recommendations. Samples were loaded into optical PCR plates and analyzed using a QuantStudio 5 Real-Time PCR System (Applied Biosystems). Thermal denaturation was monitored over a temperature range of 25°C to 99°C with a ramp rate of 0.05°C/s. Fluorescence intensity was recorded throughout the assay, and melting temperature (Tm) was calculated from the first derivative of the fluorescence curves using QuantStudio™ Design and Analysis software v.1.3.1.

T cell isolation from human blood

Whole blood was collected from healthy human volunteers into sterile blood collection bags containing the anticoagulant EDTA (Invitrogen, Cat: 15575-038). Total blood was diluted in a 1:2 ratio with Phosphate-buffer saline (PBS). Diluted blood was layered over Ficoll solution (Pharmacia Biotech, Cat: 17-0840-02) in SepMate-50 tubes (StemCell, Cat: 85450). Density centrifugation was used to remove red blood cells and the supernatant containing PBMCs were transferred into 50mL conical tubes and washed twice with PBS to remove remaining Ficoll solution and platelets. The recovered PBMCs were counted.

CD3^+^ T cells were purified by negative selection using human T cell isolation kit (StemCell, Cat: 17951) following manufacturer’s instructions. The purity of CD3^+^ T cells was assessed and found to be ≥ 85%. Purified CD3^+^ T cells were resuspended in T cell medium which included X-VIVO 15 serum-free hematopoietic cell media (Lonza, Cat: 04-418Q) supplemented with 2mM MgCl_2_ and 100nM retinoic acid (Thermo Fisher Scientific, Cat: 044540.77) which increases integrin 𝛼4$\beta$7 expression on stimulated T cells (Girard et al., 2024; Vimonpatranon et al., 2023).

*In vitro* activation of human T cells

MAdCAM-1 co-stimulation assays were performed as previously described (Sparkes et al., 2025; Vimonpatranon et al., 2023). Briefly, flat bottom tissue culture plates (Corning, Cat: 0720090) were coated overnight at 4°C with 200ng of anti-CD3 (BioLegend, Cat: 317326) + human MAdCAM-1.Fc (BioLegend, Cat: 797906) in PBS. The plates were washed with PBS, followed by culture medium equilibration. 7A2-IgG4-IL22 (50nM) or an IgG4 isotype control (BioLegend, Cat: 403701) was added to coated wells prior to the addition of 100,000 CFSE-labeled (Invitrogen, Cat: C34554) CD3+ T cells (final volume of 200μL per well) to assess the blocking ability of the bispecific. For early T cell activation analysis, one set of wells was incubated for 24 hours, after which cells were collected and stained for flow cytometry analysis of activation markers. A separate set of wells was incubated for 4 days to evaluate T cell phenotype and cytokine production. Following the 4-day incubation, supernatants were collected for cytokine analysis, and cells were harvested for flow cytometry staining.

Flow cytometry analysis of T cell activation

Viable human T cells were stained with antibodies for 30 minutes at 4°C. For Day 1 activation analysis, cells were stained with anti-CD4-PE/Cy7, 0.5:100 (BioLegend, Cat: 317411); anti-CD8-AF647, 1:100 (BioLegend, Cat: 301022); and anti-CD69-APC/Cy7, 1:100 (BioLegend, Cat:310913). For Day 4 phenotypic analysis, cells were stained with anti-CD4-PE/Cy7, 0.5:100; anti-CD8-AF647, 1:100; anti-CD27-PE/Dazzle, 1:100 (BioLegend, Cat: 302844); anti-CD45RA-APC/Cy7, 0.5:100 (BioLegend, Cat: 304128); and anti-CD25-AF700, 1:100 (BioLegend, Cat: 302622). All antibodies were diluted in PBS + 1% BSA + 0.1% sodium azide. Cells were washed, fixed and stored in PBS + 1% Paraformaldehyde (BD Biosciences, Cat: 554655). Cell populations were detected using a BD FACSSymphony A3 (BD Biosciences) cytometer (CCSM at SRI; Sunnybrook Health Sciences Center, Toronto, ON, Canada) and analyzed using FlowJo software (version XXXXXX).

Cell culture

Human COLO205 colorectal epithelial cells were used in this study. COLO205 cells were maintained in RPMI-1640 medium (Wisent, Cat: 350-000-CL) supplemented with 10% FBS (Wisent, Cat: 080450), 100 U/ml penicillin, and 100 μg/ml of streptomycin (Wisent, Cat: 450-201-EL). Cells were cultured at 37^ο^C in a humified incubator containing 95% air and 5% CO_2_ according to standard culture conditions.

Recombinant IL-22 and 7A2-IgG4-IL22 stimulation assay

COLO205 cells were seeded at a density of 75,000 cells/ cm^2^ into 48- and 96- well flat bottom tissue culture plates to assess human IL-22-mediated signaling. COLO205 cells were allowed to adhere overnight prior to stimulation. Cells were washed once with PBS prior to treatment and then stimulated with recombinant human IL-22 (50 ng/mL) (BioLegend, Cat: 571304) or molar-matched concentrations of either 7A2-IgG4-IL22 or an IgG4 isotype control (BioLegend, Cat: 403701). As a control, a five-fold molar excess of human IL-22BP was also used to inhibit IL-22 signaling, (BioLegend, Cat: 766502) . This inhibitor was added to designated conditions containing either IL-22 or the bispecific 7A2-IgG4-IL22. All stimulations were performed at 37^ο^C under standard culture conditions for each time point. Experimental readouts were performed using distinct plate formats: 48-well plates were used for qPCR analysis of IL22-responsive gene expression; and 96-well plates were used for collecting supernatants for cytokine quantification.

RNA isolation and reverse transcription polymerase chain reaction (RT-PCR, qPCR)

Gene transcript levels were determined by qPCR. COLO205 cells total RNA was extracted using the TRIzol-chloroform method following standard phenol-chloroform phase separation. Briefly, cells were lyzed in TRIzol reagent (Invitrogen, Cat: 15596018) and chloroform (Caledon, Cat: 3000-1) was added to induce phase separation. After centrifugation, the aqueous phase containing RNA was collected, then precipitated with isopropanol, washed with 75% ethanol, resuspended in ultrapure water and quantified using a NanoDrop 2000 (ThermoFisher). RNA was converted to cDNA by reverse transcription using the High-Capacity cDNA Reverse Transcription Kit (ThermoFisher, Cat: 4387406) following the manufacturer’s instructions. Gene expression was quantified with pre-designed TaqMan Fast Advanced Master Mix (Applied Biosystems, Cat: 4444557) on a QuantStudio 5 thermocycler (Applied Biosystems) to investigate markers of IL22-signaling. Gene expression was then normalized using the 2-ΔΔCq method relative to the expression of the housekeeping gene, GAPDH. Primers used include GAPDH (Hs02758991_g1), REG3A (Hs00170171_m1), S100A8 (Hs00375264_g1), CXCL1 (Hs00236937_m1), IL10 (Hs00961619_m1), and CLDN2 (Hs01568822_m1)

Cytokine release assay

Cytokine protein levels in cell culture supernatants were quantified using the LEGENDplex^TM^ human inflammation panel 1 (BioLegend, Cat: 740809) according to the manufacturer’s instructions. For T cell activation assays, supernatants collected after 4 days of stimulation were analyzed for tumor necrosis factor (TNF) α, and interferon gamma (IFN𝛾). In addition, supernatants collected following a 48-hour stimulation of COLO205 cells were analyzed for IL-8 and IL-10 expression using the same panel. The samples were read using the BD FACSSymphony A3 (BD Biosciences) cytometer, and the data were analyzed using the cloud-based LEGENDplex^TM^ Data Analysis Software (BioLegend).

MTT Assay

Cell metabolic activity was assessed using the MTT Cell Proliferation Assay Kit (Thermo Fisher Scientific, Cat: V13154) according to the manufacturer’s instructions. The MTT assay quantifies the reduction of the tetrazolium salt MTT by metabolically active cells into insoluble purple formazan crystals through cellular oxidoreductase enzymes, providing an indirect measure of cell metabolic activity and viability. Briefly, COLO205 cells were seeded in 96-well plates and stimulated for 24 hours with recombinant human IL-22 (50 ng/mL) (BioLegend, Cat: 571304) or molar-matched concentrations of either 7A2-IgG4-IL22 or an IgG4 isotype control (BioLegend, Cat: 403701). As a control, a five-fold molar excess of human IL-22BP was also used to inhibit IL-22 signaling, (BioLegend, Cat: 766502). Following stimulation, 100μL of culture supernatant was removed from each well, and 10μL of MTT reagent was added directly to the remaining culture. Plates were then transferred to a 37^ο^C incubator for 4 hours to allow the formation of intracellular formazan crystals. The plates were centrifuged and the culture medium aspirated and replaced with dimethyl sulfoxide (DMSO) to solubilize the formazan crystals. Absorbance was measured at 570nm using a Synergy H1 microplate reader. Optical density values were blank-adjusted prior to analysis and normalized to unstimulated controls. Average optical density measurements ± SD were derived from four independent experiments (n=4).
